# Preserved Medial Temporal Lobe Flexibility Predicts Memory Generalization Only in the Context of Good Sleep Quality among Older African Americans

**DOI:** 10.64898/2026.06.15.26355704

**Authors:** Payton White, Miray Budak, Soodeh Moallemian, Bernadette A. Fausto, Mark A. Gluck

**Author notes:** Corresponding author; email: Payton White. ***e-mail addresses of Authors***.

## Abstract

**Objectives:** Poor sleep quality is a risk factor for Alzheimer’s disease (AD). Older African Americans experience disproportionately high rates of sleep disturbance and AD. Medial temporal lobe (MTL) flexibility reflects dynamic neural reorganization and may be a marker of generalization performance. This study examined whether sleep quality moderates the association between MTL flexibility and memory generalization.

**Methods:** Fifty older African Americans (MeanAge=69.7±6.21 years; 80% women) underwent rs-fMRI to quantify MTL flexibility, Rutgers Acquired Equivalence Task for memory generalization, and Pittsburgh Sleep Quality Index for sleep quality.

**Results:** Greater MTL flexibility was associated with better generalization (r=0.367, p=.017). Good sleepers showed higher MTL flexibility (F_(1,44)_=8.11, ηp^2^=.156, p=.007) and superior generalization (F_(1,46)_= 12.33, ηp^2^=.211, p=.001). Sleep quality significantly moderated the MTL flexibility–generalization relationship (β=−1.519, p=.012).

**Conclusions:** Preserved MTL flexibility may confer generalization only in good sleepers, suggesting that sleep disturbance may disrupt the MTL neural resilience among older African Americans.

**WHAT WAS KNOWN:** Prior research has established that poor sleep quality is associated with increased Alzheimer’s disease (AD) risk, hippocampal dysfunction, and impaired memory. Studies also show that medial temporal lobe (MTL) network dynamics, including flexibility, support key features of cognition including memory generalization. However, limited work has examined how sleep quality interacts with neural flexibility to influence cognition, particularly in older African Americans, a population at elevated risk for both sleep disturbance and AD.

**WHAT THIS STUDY ADDS:** This study demonstrates that sleep quality significantly moderates the relationship between MTL flexibility and memory generalization. While greater MTL flexibility is associated with better generalization, this benefit is evident only among individuals with good sleep quality. These findings suggest that sleep disruption may impair the functional expression of neural resilience mechanisms, even when underlying network flexibility is preserved.

## INTRODUCTION

Sleep disruption is increasingly recognized as both an early feature and a contributing factor in Alzheimer’s disease (AD), reflecting a bidirectional relationship between sleep quality and neurodegeneration.^1^ Poor sleep characterized by reduced efficiency, increased fragmentation, and diminished slow-wave activity^2,3^ is associated with amyloid and tau pathology, neuroinflammation, and impaired hippocampal function.^4,5^ Older African Americans experience a disproportionate burden of AD as well as higher rates of sleep disturbances,^6,7^ yet remain underrepresented in mechanistic neuroimaging research.^8^ Understanding neural pathways through which sleep influences cognition in this high-risk population is essential for precision prevention strategies.

The medial temporal lobe (MTL), central to learning and episodic memory, is among the earliest structures to accumulate AD pathology.^9^ The hippocampus within the MTL supports memory generalization the ability to apply learned associations to novel contexts a function that declines with aging and early disease-related changes.^10^ Dynamic network connectivity (“flexibility”) within the MTL, assessed via resting-state fMRI (rs-fMRI), reflects time-varying reconfiguration of functional connectivity and has emerged as a marker of neural resilience.^11^

Sleep supports hippocampal plasticity and memory consolidation through synaptic homeostasis, memory replay, and slow-wave activity.^13^ Disruption of these processes may compromise neural network efficiency and contribute to early cognitive vulnerability.^14^Subjective sleep quality is commonly captured with validated tools such as the Pittsburgh Sleep Quality Index (PSQI).^15^ Despite converging evidence linking sleep, hippocampal function, and cognition, it remains unknown whether sleep quality influences the extent to which preserved MTL network dynamics translate into effective generalization performance.

We hypothesized that (1) greater MTL flexibility would be associated with better generalization, (2) individuals with better sleep quality would exhibit higher MTL flexibility and generalization performance, and sleep quality would moderate the MTL flexibility–generalization association.

## METHODS

### Participants

Fifty cognitively unimpaired older African Americans from Pathways to Healthy Aging in African Americans—a longitudinal study at Rutgers University–Newark were enrolled. Participants were aged ≥60, without neurodegenerative diagnoses, dementia medications, learning disabilities, alcohol/substance misuse, or recent general anesthesia. The study was approved by the Rutgers IRB and adhered to the Declaration of Helsinki.

### Measures

Subjective sleep quality was assessed with the PSQI.^15^ A total score >5 indicated poor sleep quality.^16^ Memory generalization was assessed with the Rutgers Acquired Equivalence Task, which requires participants to apply learned stimulus-response associations to novel transfer trials.^10^ Global cognition was characterized with the Montreal Cognitive Assessment (MoCA); cutoffs were adjusted for race/ethnicity and education.^14^

### MRI Acquisition and Analysis

MRI data were acquired at the Rutgers University Brain Imaging Center on a 3T Siemens Prisma scanner. Resting-state fMRI (rs-fMRI) data were preprocessed using AFNI’s afni_proc.py pipeline, including despiking, slice-timing correction, motion correction, spatial smoothing (2 mm FWHM), and nuisance regression. Dynamic functional connectivity was computed within seven MTL regions of interest (subiculum, CA1, DG/CA3, perirhinal cortex, parahippocampal cortex, posteromedial and anterolateral entorhinal cortex) across nine non-overlapping 50-TR windows using magnitude-squared spectral coherence (0.06–0.12 Hz). Multilayer modularity optimization yielded node flexibility scores; mean flexibility across all ROIs served as the primary neural outcome.

### Statistical Analysis

All statistical analyses were conducted using Jamovi (version 2.7.31.0). Data were screened for normality and outliers through inspection of descriptive statistics, distribution plots, and residual diagnostics. Missing data were minimal (<5%) and handled using available-case analyses, such that each statistical model included all participants with non-missing values for the variables entered in that model; no data imputation was applied.

Pearson correlation analyses were performed to examine the association between MTL flexibility and generalization. Group differences in MTL flexibility and generalization between participants with good and poor sleep quality were evaluated using general linear models. To test whether sleep quality moderated the association between MTL flexibility and generalization, a linear regression model including the interaction term (MTL flexibility × sleep quality) was conducted. Sleep quality was entered as a categorical predictor, and MTL flexibility was treated as a continuous variable. All models were adjusted for age, sex, and years of education. Although MoCA scores differed significantly between sleep groups, MoCA was not included as a covariate because all participants fell within the cognitively normal range based on education- and ethnicity-adjusted cutoffs. Importantly, sensitivity analyses including MoCA as a covariate yielded the same pattern of results, indicating that the findings were not driven by global cognitive differences. Effect sizes (ηp^2^ for group comparisons; standardized β coefficients for regression models), model fit indices (R^2^), and two-tailed p-values are reported. Statistical significance was set at p < .05.

## RESULTS

### Participant Characteristics

The sample included 50 older African Americans (mean age = 67.70 ± 6.21 years; range = 60–83), of whom 80% were women. Participants averaged 14.00 ± 2.30 years of education and scored within the cognitively unimpaired range on the MoCA (23.60 ± 2.46). Based on PSQI criteria, 39 participants were classified as poor sleepers and 11 as good sleepers. Groups did not differ in age (t(47) = 0.47, p = .642) or education (t(46) = −1.02, p = .313), though the poor sleep group had a higher proportion of women (t(47) = −2.43, p = .019) and higher MoCA scores (t(46) = 2.19, p = .034; all within the unimpaired range). Demographic characteristics are summarized in Table 1.

**Table 1.** Distribution of demographic data.

| | | All Cohort Mean ± SD | Min/Max | Good Sleep Quality Group (N = 11 ) | Poor Sleep Quality Group (N =39 ) | Statistics |
| --- | --- | --- | --- | --- | --- | --- |
| Age (years) | | 67.70 ± 6.21 | 60 / 83 | 68.90 ± 5.87 | 69.90 ± 6.39 | t(47)= 0.47, p= .642 |
| Sex N (%) | Women | 40 (80) | -- | 6 (12) | 34 (68) | t(47)= -2.43, p = .019 |
|  | Men | 10 (20) | -- | 5 (10) | 5 (10) |  |
| Education (years) | | 14.00 ± 2.30 | 7 / 18 | 14.5 ± 1.69 | 13.9 ± 2.38 | t(46)= -1.02, p = .313 |
| MoCA | | 23.60 ± 2.46 | 19 / 29 | 22.9 ± 2.49 | 23.8 ± 2.31 | t(47)= 2.19, p = .034 |
| MTL Flexibility | | 0.381 ± 0.223 | 0.00 / 1.00 | 0.516 ± 0.183 | 0.343 ± 0.221 | t(48)= -2.365, p= .022 |
| Rutgers Acquired Equivalence Task | | 0.599 ± 0.269 | 0.00 / 1.00 | 0.682 ± 0.304 | 0.575 ± 0.262 | t(25)= -0.848, p= .405 |

Max = Maximum; Min = Minimum; MoCA = Montreal Cognitive Assessment; N = Number; PSQI = Pittsburgh Sleep Quality Index; SD = Standard Deviation.

### Effect of Sleep Quality on MTL Flexibility and Memory Generalization

Participants with good sleep quality showed significantly higher MTL flexibility (F(1,44) = 8.11, p = .007, ηp^2^ = .156; Figure 1a) and greater generalization accuracy (F(1,46) = 12.33, p = .001, ηp^2^ = .211; Figure 1b) compared to those with poor sleep quality.

**Figure 1.**
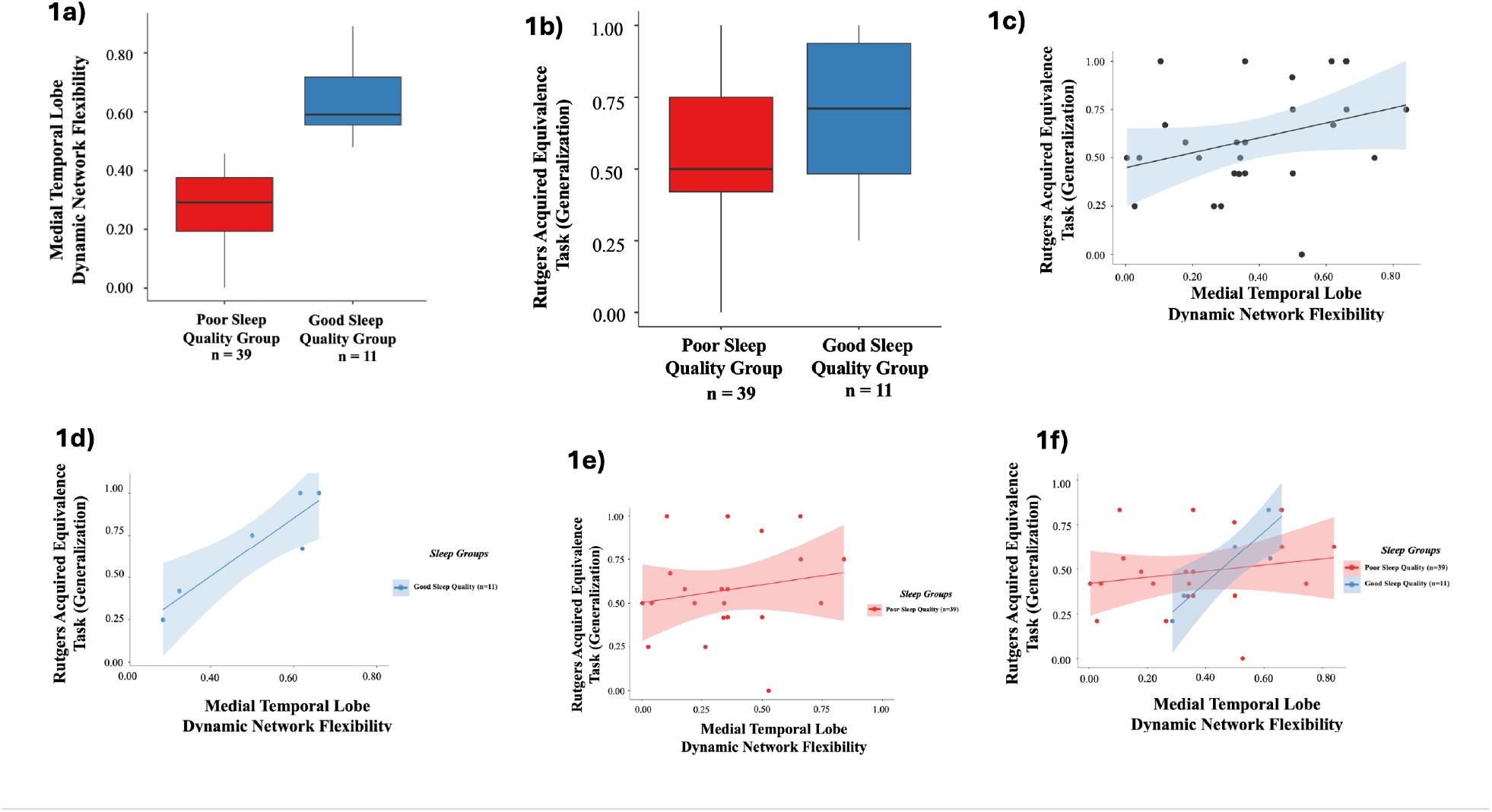
A) Participants with good sleep quality exhibit higher MTL flexibility compared to participants with poor sleep quality. **B)** Participants with good sleep quality demonstrate superior memory generalization compared to participants with poor sleep quality. **C)** Greater MTL flexibility is associated with better memory generalization. **D-F)** Sleep quality moderates the association between MTL flexibility and memory generalization.

### Relationship Between MTL Flexibility and Memory Generalization

Across the full sample, greater MTL flexibility was significantly associated with better generalization (r = .367, p = .017; Figure 1c), indicating that increased time-varying neural reconfiguration within the MTL relates to enhanced cognitive flexibility.

### Moderation Effect of Sleep Quality

Sleep quality significantly moderated the MTL flexibility–generalization association (β = −1.519, p = .012; Figure 1f). Simple slopes analysis revealed that greater MTL flexibility predicted better generalization only among good sleepers (Figure 1d), whereas no significant relationship was observed among poor sleepers (p > .05; Figure 1e).

## DISCUSSION

This study demonstrates that sleep quality moderates the relationship between MTL dynamic network flexibility and memory generalization in cognitively unimpaired older African Americans. Although greater MTL flexibility was broadly associated with better generalization, this benefit was present only among individuals with good sleep quality.

MTL flexibility alone may be insufficient to support adaptive memory function.^20^ Even poor sleepers with relatively higher MTL flexibility showed variable and lower generalization performance, suggesting that sleep disruption may interfere with the effective utilization of neural capacity rather than simply reducing it.^17^ Sleep may thus determine whether preserved neural dynamics are translated into stable behavioral representations; their decoupling in poor sleepers may reflect reduced efficiency in linking neural organization to cognition.^18^

A plausible mechanism involves sleep-dependent hippocampal consolidation.^19^ Slow-wave sleep supports memory replay, synaptic homeostasis, and hippocampal–cortical communication,^20^ facilitating integration of new information into existing knowledge structures essential for generalization. Sleep disruption may yield less stable memory representations,^21^ such that higher MTL flexibility during poor sleep reflects neural variability without corresponding gains in functional integration.

These findings are consistent with work linking poor sleep to hippocampal dysfunction and elevated AD risk.^22,23^ The present results extend this literature by suggesting that sleep quality may influence not only neural function and cognition independently, but also the relationship between them.^24^ Older African Americans are disproportionately affected by both sleep disturbance and AD, highlighting sleep as a modifiable target in this high-risk population.

Limitations include reliance on subjective sleep measurement, a cross-sectional design, and absence of biomarkers. Future studies should incorporate objective sleep measures, longitudinal designs, and plasma biomarkers (e.g., p-tau, Aβ) to clarify causal pathways and disease relevance.

## CONCLUSION

Preserved MTL flexibility may not be sufficient to support cognitive performance without adequate sleep. Sleep quality may influence the extent to which neural capacity is translated into effective behavior, highlighting it as a potentially modifiable factor for cognitive resilience in aging and AD prevention, particularly among older African Americans.

## Data Availability

All data produced in the present study are available upon reasonable request to the authors.

## Declaration of the Use of Generative AI and AI-Assisted Technologies

The authors declare that no generative artificial intelligence (AI) or AI-assisted technologies were used in the writing of this manuscript or in the creation of figures, images, or artwork.

## Author Contribution Payton White

Writing – review & editing, Writing – original draft, Visualization, Methodology, Formal analysis, Conceptualization. **Miray Budak:** Writing – review & editing, Writing – original draft, Supervision, Methodology. **Soodeh Moallemian:** Writing – review & editing, Conceptualization. **Bernadette A. Fausto:** Writing – review & editing, Supervision, Conceptualization. **Mark A. Gluck:** Writing – review & editing, Supervision, Resources, Methodology, Funding acquisition, Conceptualization.

## Data Sharing

De-identified data supporting the findings of this study are available from the corresponding author upon reasonable request and subject to institutional approval and participant confidentiality requirements.

## Acknowledgments

We are grateful for the feedback and shared insights from our Community Brain Health Educators and Outreach Team: Glenda Wright, Delores Hammonds, Jerome Perkins, Louches Powell, Reverend Glenn Wilson, and Catherine Willis. We are indebted to the thousands of community members who have participated in our brain health events since 2006, and to over 450 community members who have enrolled, to date, as VIPs (Very Important Participants) in our Aging & Brain Health Alliance study.

## REFERENCES

1. Wang C, Holtzman DM. Bidirectional relationship between sleep and Alzheimer’s disease: role of amyloid, tau, and other factors. Neuropsychopharmacology. 2020;45(1):1. doi:10.1038/s41386-019-0478-5

2. Varga AW, Wohlleber ME, Giménez S, et al. Reduced Slow-Wave Sleep Is Associated with High Cerebrospinal Fluid Aβ42 Levels in Cognitively Normal Elderly. Sleep. 2016;39(11):2041–2048. doi:10.5665/sleep.6240

3. Ju YES, Ooms SJ, Sutphen C, et al. Slow wave sleep disruption increases cerebrospinal fluid amyloid-β levels. Brain. 2017;140(8):2104–2111. doi:10.1093/brain/awx148

4. Shen Y, Lv Q kun, Xie W ye, et al. Circadian disruption and sleep disorders in neurodegeneration. Transl Neurodegener. 2023;12:8. doi:10.1186/s40035-023-00340-6

5. Mander BA, Dave A, Lui KK, et al. Inflammation, tau pathology, and synaptic integrity associated with sleep spindles and memory prior to β-amyloid positivity. Sleep. 2022;45(9):zsac135. doi:10.1093/sleep/zsac135

6. Gillis C, Montenigro P, Nejati M, Maserejian N. Estimating prevalence of early Alzheimer’s disease in the United States, accounting for racial and ethnic diversity. doi:10.1002/alz.12822

7. Jean-Louis G, Magai CM, Cohen CI, et al. Ethnic Differences in Self-Reported Sleep Problems in Older Adults. Sleep. 2001;24(8):926–933. doi:10.1093/sleep/24.8.926

8. Lim AC, Barnes LL, Weissberger GH, et al. Quantification of race/ethnicity representation in Alzheimer’s disease neuroimaging research in the USA: a systematic review. Commun Med. 2023;3(1):101. doi:10.1038/s43856-023-00333-6

9. Aksman LM, Oxtoby NP, Scelsi MA, et al. A data-driven study of Alzheimer’s disease related amyloid and tau pathology progression. Brain. 2023;146(12):4935–4948. doi:10.1093/brain/awad232

10. Myers CE, Kluger A, Golomb J, et al. Hippocampal Atrophy Disrupts Transfer Generalization in Nondemented Elderly. J Geriatr Psychiatry Neurol. 2002;15(2):82–90. doi:10.1177/089198870201500206

11. Sperling RA, Dickerson BC, Pihlajamaki M, et al. Functional Alterations in Memory Networks in Early Alzheimer’s Disease. NeuroMolecular Med. 2010;12(1):27–43. doi:10.1007/s12017-009-8109-7

12. Brodt S, Inostroza M, Niethard N, Born J. Sleep—A brain-state serving systems memory consolidation. Neuron. 2023;111(7):1050–1075. doi:10.1016/j.neuron.2023.03.005

13. Tononi G, Cirelli C. Sleep and the Price of Plasticity: From Synaptic and Cellular Homeostasis to Memory Consolidation and Integration. Neuron. 2014;81(1):12–34. doi:10.1016/j.neuron.2013.12.025

14. Milani SA, Marsiske M, Cottler LB, Chen X, Striley CW. Optimal cutoffs for the Montreal Cognitive Assessment vary by race and ethnicity. Alzheimers Dement Diagn Assess Dis Monit. 2018;10(1):773–781. doi:10.1016/j.dadm.2018.09.003

15. Buysse DJ, Reynolds CF, Monk TH, Berman SR, Kupfer DJ. The Pittsburgh Sleep Quality Index: a new instrument for psychiatric practice and research. Psychiatry Res. 1989;28(2):193–213. doi:10.1016/0165-1781(89)90047-4

16. Buysse DJ, Reynolds CF III, Monk TH, et al. Quantification of Subjective Sleep Quality in Healthy Elderly Men and Women Using the Pittsburgh Sleep Quality Index (PSQI). Sleep. 1991;14(4):331–338. doi:10.1093/sleep/14.4.331

17. Boonstra TW, Stins JF, Daffertshofer A, Beek PJ. Effects of sleep deprivation on neural functioning: an integrative review. Cell Mol Life Sci CMLS. 2007;64(7-8):934. doi:10.1007/s00018-007-6457-8

18. Tibon R, Tsvetanov KA. The “Neural Shift” of Sleep Quality and Cognitive Aging: A Resting-State MEG Study of Transient Neural Dynamics. Front Aging Neurosci. 2022;13:746236. doi:10.3389/fnagi.2021.746236

19. Havekes R, Abel T. The tired hippocampus: The molecular impact of sleep deprivation on hippocampal function. Curr Opin Neurobiol. 2017;44:13–19. doi:10.1016/j.conb.2017.02.005

20. Whitmore NW, Yamazaki EM, Paller KA. Targeted memory reactivation with sleep disruption does not weaken week-old memories. NPJ Sci Learn. 2024;9:64. doi:10.1038/s41539-024-00276-0

21. Zhou X, Lauharatanahirun N, Thurman SM, et al. Flexibility of Brain Networks May Curtail Cognitive Consequences of Poor Sleep. Hum Brain Mapp. 2025;46(14):e70362. doi:10.1002/hbm.70362

22. Li K, Luo X, Zeng Q, et al. Interactions between sleep disturbances and Alzheimer’s disease on brain function: a preliminary study combining the static and dynamic functional MRI. Sci Rep. 2019;9(1):19064. doi:10.1038/s41598-019-55452-9

23. Kent BA, Mistlberger RE. Sleep and hippocampal neurogenesis: Implications for Alzheimer’s disease. Front Neuroendocrinol. 2017;45:35–52. doi:10.1016/j.yfrne.2017.02.004

24. Hyndych A, El-Abassi R, Mader EC. The Role of Sleep and the Effects of Sleep Loss on Cognitive, Affective, and Behavioral Processes. Cureus. 17(5):e84232. doi:10.7759/cureus.84232

